# *PBRM1* loss-of-function mutations and response to immune checkpoint blockade in clear cell renal cell carcinoma

**DOI:** 10.1101/2020.10.30.20222356

**Authors:** Jake Conway, Amaro Taylor-Weiner, David Braun, Ziad Bakouny, Toni K. Choueiri, Eliezer M. Van Allen

## Abstract

The biological role of *PBRM1* loss-of-function (LOF) mutations mediating response to immune checkpoint blockade (ICB) in clear cell renal cell carcinoma (ccRCC) remains an area of active investigation. A recent study (hereafter, ‘the MSK study’) evaluated this hypothesis in a clinically heterogeneous single institution retrospective ccRCC cohort using targeted sequencing and did not find a univariable statistically significant association for *PBRM1* LOF and time-to-treatment failure (TTF), but did report a significant association between continuous tumor mutational burden (TMB) and ICB TTF. We re-analyzed this cohort to (i) match the methodology from previous studies and (ii) account for differences in cohort development, statistical approaches, mutation classifications, and outcome measurements. Univariable analysis of the *PBRM1* LOF mutation hypothesis, when performed analogously to prior studies, demonstrated a modest statistically significant association with TTF on ICB (p = 0.047; HR = 0.7, 95% CI = 0.49 - 0.99). By contrast, when using the different methodology presented in the MSK study, statistical modeling demonstrated that there was insufficient power to detect this association given the altered classification scheme and modest effect size. In addition, after appropriately normalizing TMB to account for anomalous outliers, TMB was not significantly associated with ICB response in ccRCC. Thus, this study provides further support for the biological association between *PBRM1* LOF and favorable clinical outcomes on ICB in specific ccRCC clinical contexts. However, we again strongly caution against interpreting that this *PBRM1* biological association, likely one of many modest mediators of tumor-immune-stromal interactions, achieves broadly generalizable predictive biomarker status for ICB in diverse clinical contexts. The *PBRM1* LOF biological association has only been shown to be modestly predictive in the post-VEGF TKI ICB-monotherapy ccRCC patient population. Given emerging data about the biological role of *PBRM1* in contributing to interwoven immune and angiogenesis programs (among many others), and the rapidly shifting therapeutic combination strategies invoking these processes in ccRCC, focused biological and predictive biomarker analyses in specific clinical settings using appropriately annotated clinical trial cohorts are necessary.

## MAIN TEXT

Two clear cell renal cell carcinoma (ccRCC) clinical trial cohorts evaluating single-agent immune checkpoint blockade (ICB) in a VEGF receptor tyrosine kinase inhibitor (VEGF TKI)-refractory patient population identified a statistically significant association between *PBRM1* loss-of-function (LOF) mutations and ICB response (1-3), whereas two trial cohorts in the first-line treatment-naive metastatic setting with different ICB-based therapeutic combinations did not observe a significant association (4, 5). Hakimi et al (hereafter, ‘the MSK study’) investigated associations of mutations in *PBRM1* and other PBAF complex members with response to immune checkpoint blockade (ICB) in a clinically heterogeneous single institution retrospective cohort of ccRCC, and generally across cancer types (6). They did not observe a significant association for *PBRM1* LOF in ccRCC in univariable analysis, nor in multivariable analysis incorporating two factors they found significantly associated with ICB response in ccRCC: tumor mutational burden (TMB) and treatment class. These observations raise key questions for the field about the biological and biomarker relevance of *PBRM1* mutations in ccRCC and cancer immunotherapy, and also more generally the interpretation of candidate molecular mediators of ICB response from biological and clinical perspectives.

To investigate the MSK study in the context of prior reports, we first assessed the univariable outcomes in ccRCC reported in this study using the processed data provided to us by the corresponding author. Given the desire to evaluate clinical outcomes that directly link to potential biological relevance of these findings, we first examined time-to-treatment failure (TTF) in this cohort (as a proxy for progression free survival [PFS]). Information on patient selection or technical quality control from sequencing data was not reported or made available for sensitivity analyses to identify potential biases. Beyond these general issues, the MSK study differed from prior studies in two key ways: 1) They chose different criteria for LOF mutations (excluding splice site mutations as LOF variants for this tumor suppressor, even though this research group and others generally considered this variant class as LOF in prior genomic or immunohistochemistry studies (7-12)); and 2) They split the control arm into two groups (*PBRM1* non-LOF that included splice site mutated patients, and *PBRM1* wild-type). As a result, power was reduced to detect a significant association between *PBRM1* LOF and ICB response by including putative LOF variants in a separated non-LOF control group and by lowering the sample size of each group.

When we re-evaluated this dataset using the previously established definition of *PBRM1* LOF mutations (1-3) to directly assess the LOF hypothesis, we observed a statistically significant association between *PBRM1* LOF and improved TTF with ICB (p = 0.047; HR = 0.7, 95% CI = 0.49 - 0.99; Fig. 1A; Methods). We noted that there were minimal differences between the statistically significant HR in our standardized analysis and the original study’s non-statistically significant HR estimate (HR = 0.7 vs 0.73), and a small difference in p-values (p = 0.047 vs p = 0.11 in our analysis and the original MSK study, respectively). Since the methodological choices in the MSK study decreased statistical power to detect the same effect size for this hypothesis, we then simulated the minimum cohort sizes necessary to observe a significant association between *PBRM1* LOF mutations and TTF, given the study population, effect size and the statistical results reported in the originating study (Methods). This simulation indicated that, with the cohort size, composition, and originating analytical setup, there was 39% power to detect a statistically significant difference (Fig. 1B), and the subgroup analyses split by line of therapy were similarly underpowered (Fig. 2A-B; Methods). This observation highlights the challenges of binary interpretation of p-values, specifically differentiating between statistically significant results (which are highly dependent on sample sizes) and either biologically or clinically significant results (13). After adjusting the PBRM1 classifications to match multiple prior studies (1, 3), their univariable results from a single institution, retrospective ccRCC cohort therefore provides further support for the significant but modest effect size association between *PBRM1* LOF mutations and ICB response.

**Figure 1:**
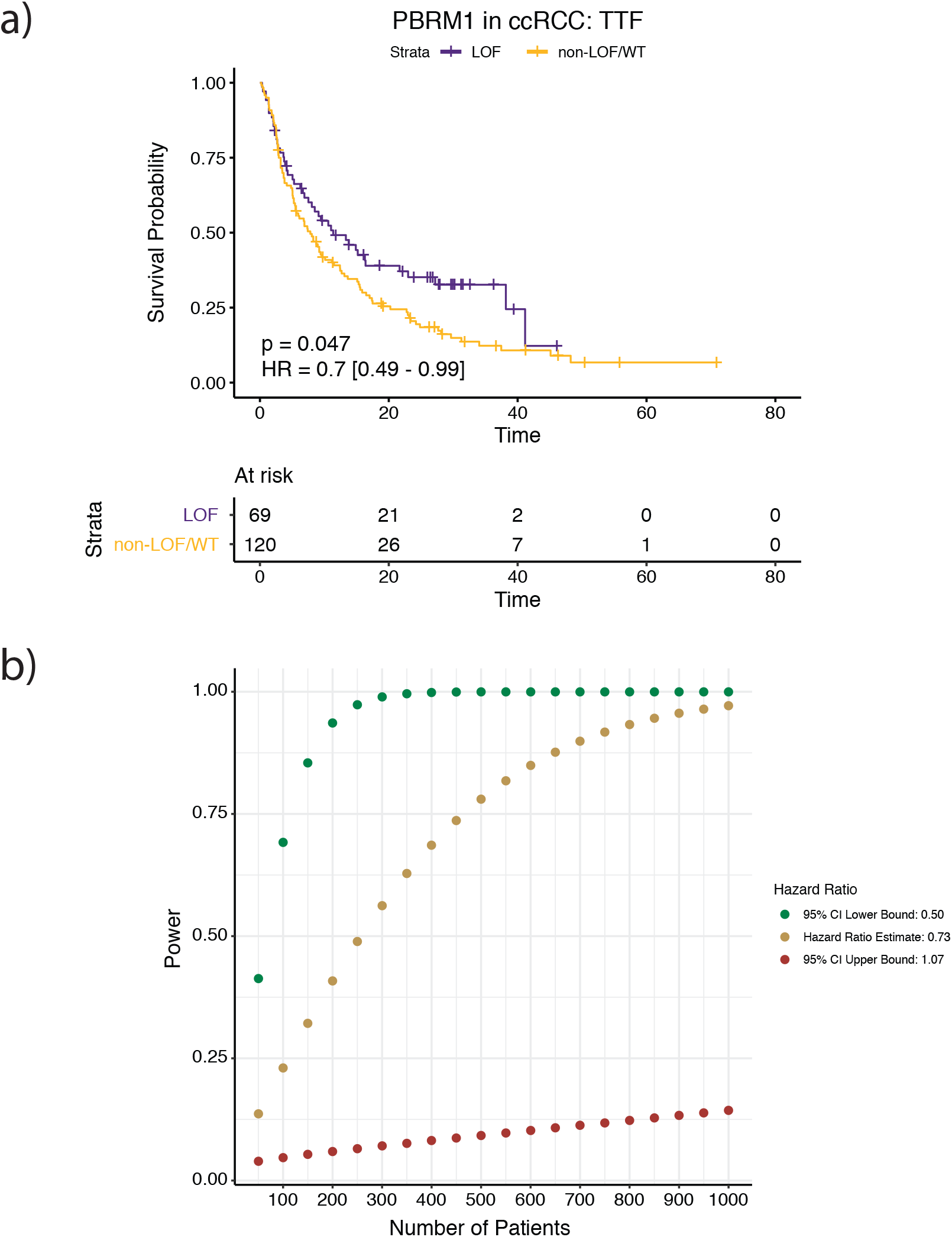
Effect of *PBRM1* classification criteria from previously published studies and univariable Cox proportional hazards power simulations using the originating study criteria. **a)** When classifying *PBRM1* alterations as LOF using the same criteria as previous studies (i.e. including splice-site mutations as LOF), and keeping the non-LOF patients as a single control arm like previous studies, there is a significant improvement in TTF for *PBRM1* LOF patients (Cox PH, p = 0.047, HR = 0.7, 95% CI = 0.49 - 0.99). **b)** Simulations showing the power to determine that *PBRM1* LOF mutations have a significantly different hazard by the number of patients included in the cohort. The proportion of *PBRM1* LOF patients used in the power calculations was 32%, the same as the originating study. The power calculations were performed under the assumptions that the true hazard ratio was the lower bound of the 95% confidence interval (green), the hazards ratio estimate (gold), and the upper bound of the 95% confidence interval (brown) from the univariable Cox proportional hazards analysis in the originating study.

**Figure 2:**
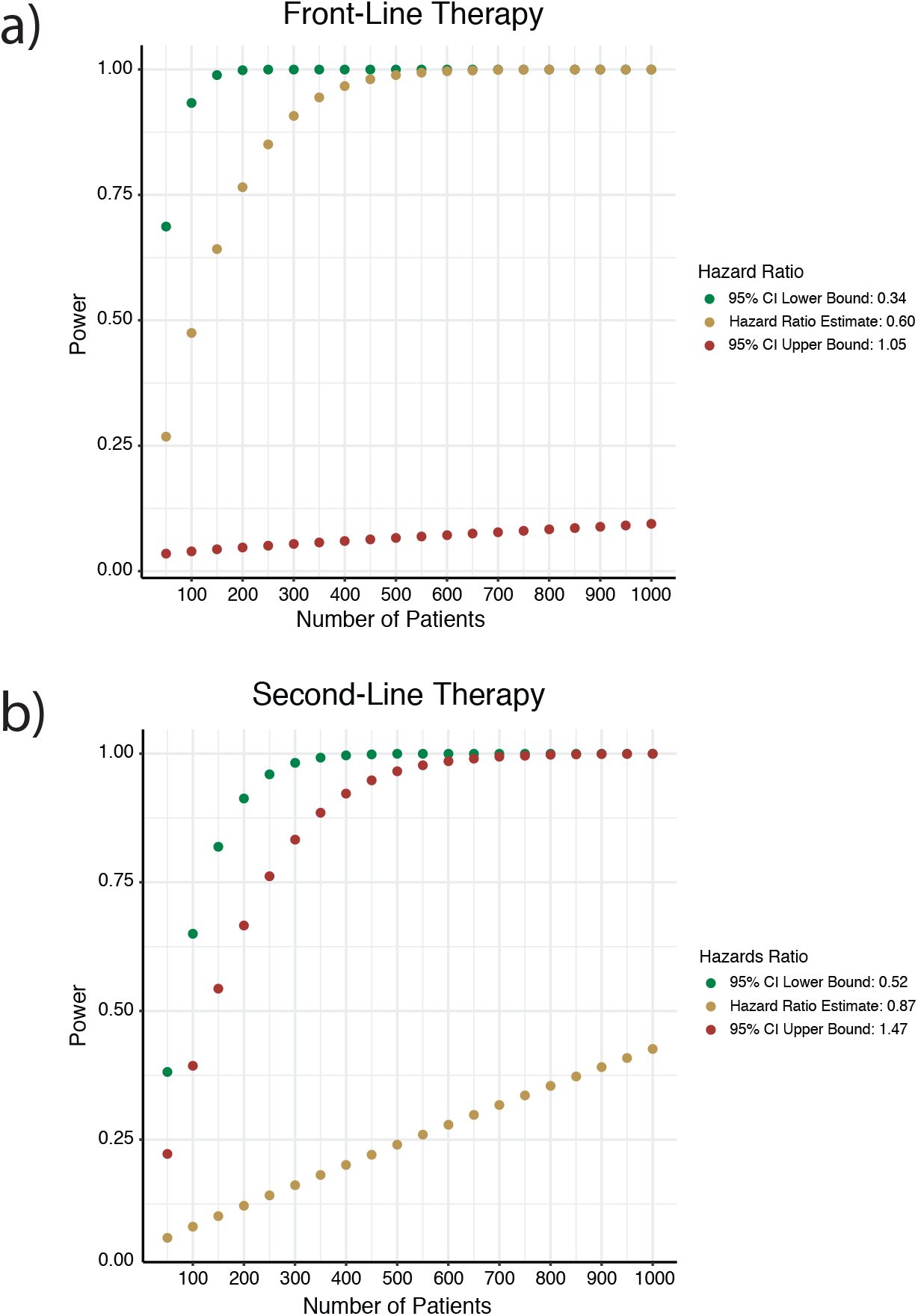
Univariable Cox proportional hazards power simulations by line of therapy using the originating study criteria. Simulations showing the power to determine that *PBRM1* LOF mutations have a significantly different hazard in the **a)** front-line setting and **b)** second-line setting by the number of patients included in the cohort. The proportion of *PBRM1* LOF patients used in the power calculations was 32%, the same as the overall *PBRM1* LOF frequency originating study. The power calculations were performed under the assumptions that the true hazard ratio was the lower bound of the 95% confidenc interval (green), the hazards ratio estimate (gold), and the upper bound of the 95% confidence interval (brown) from univariable Cox proportional hazards analysis on patients from **a)** front-line and **b)** second-line settings, respectively.

Regarding the overall survival (OS) univariable analyses, the reported OS and time-to-treatment failure (TTF) findings for *PBRM1* in ccRCC (n = 189 patients) yielded opposing hazard ratios, and the median OS for this advanced ccRCC cohort was 68.2 months. This median OS for the patients in this study, which was a mix of first-line and >= 2 line metastatic ccRCC patients, far exceeded the median OS of any known metastatic ccRCC clinical trial cohort in either first-line or >= second line treatment settings (e.g. one modern clinical trial of first-line treatment-naive ccRCC patients treated with ICBs reported a median OS of 47 months (14), and a second trial in >= 2 line setting reported median OS of 25 months (15)). In the MSK study, OS was defined from ccRCC diagnosis at any stage, rather than the standard definition of time from start of ICB therapy for metastatic ccRCC (A. Hakimi, personal communication, September 10, 2020), rendering the reported OS challenging to interpret. This issue illustrates a general issue in retrospective single institution molecular analyses, whereby unaccountable biases (e.g. whether drug class impacted censoring, timing of biopsy used for sequencing relative to treatment exposures, among many others) may influence interpretation of results in unknowable ways that can be circumvented through analysis of homogeneously collected tumor samples from well-defined cohorts within specific clinical settings, e.g. clinical trial cohorts (preferably randomized clinical trials).

While the original MSK study identified no association with *PBRM1* LOF, they reported a significant univariable association between continuous TMB and ICB response in ccRCC. This observation would have major clinical implications for ccRCC if validated, but contrasts findings from prior studies in ccRCC (1, 2, 4, 5, 8, 16). However, this TMB analysis had two key issues. First, their reported significant effect of TMB as a continuous predictor is driven primarily by irregularities in the TMB data and the outcomes of a handful of outliers, rather than a linear relationship to TTF. Specifically, ccRCC patients in this study had a higher median TMB than previously reported in metastatic ccRCC (3.9 mut/Mb) (2, 4), 7 patients (3.7%) had TMB = 0 mutations/Mb (noting unknown technical quality control, and expecting ccRCC to have > 0 mutations with expanded sequencing) and 3 patients (1.6%) had TMB > 10 mutations/Mb (Fig. 3A). TMB inflation is expected since the authors are reporting TMB derived from a targeted driver gene panel; however, since there were no harmonization efforts included in the study, it is impossible to distinguish between the effect of TMB versus the number of driver events (one of which is *PBRM1*). To demonstrate this, we compared univariable models including TMB or log-transformed TMB (Fig. 3B). Log transformation is routinely applied in such TMB analyses to bring outliers closer to the rest of the data distribution, reducing the effect of outliers on the estimated model coefficients (16-18). After log-transforming TMB, there was no significant association with TTF (log-transformed TMB: Cox PH, p = 0.16, HR = 0.85, 95% CI = 0.68 - 1.07; Methods). Further, because the observed effect of TMB may actually be driven by having a higher number of *PBRM1* mutations, we re-assessed the relationship between TMB and TTF in samples without *PBRM1* LOF mutations. When categorizing TMB by tertile and completely removing the effect of *PBRM1* LOF mutations on TTF, there was no difference in the TTF between samples in the highest and lowest TMB tertiles (p = 0.52, log-rank test; Methods; Fig. 3C).

**Figure 3:**
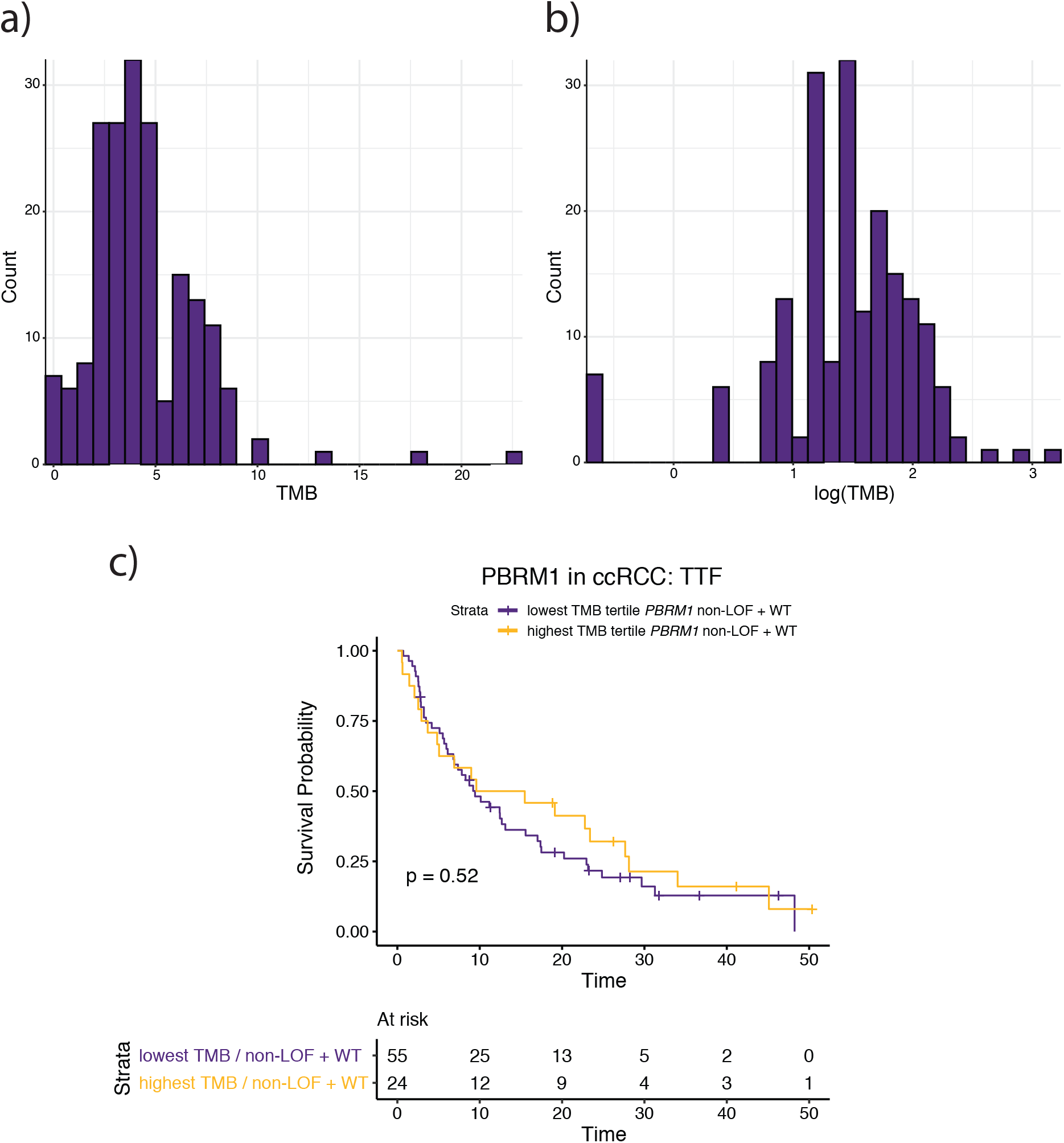
The TMB association in originating study is driven by outliers and TMB is not associated with ICB response in ccRCC. **a)** The distribution of TMB in the originating study cohort (n = 189). A total of 7 samples (3.7%) have a TMB of 0 and 3 samples (1.6%) have a TMB > 10 mutations/Mb (occurs at a rate of < 1% in ccRCC). **b)** The distribution of log-transformed TMB from the originating study cohort (n = 189). Log-transformation reduces the effect of outliers on the data, and after log-transformation TMB is no longer associated with ICB response (Cox PH, p = 0.16, HR = 0.85, 95% CI = 0.68 - 1.07). **c)** TTF survival curves between the lowest and highest TMB tertiles filtered down to non-LOF and WT *PBRM1* samples.

The TMB analysis issue was further compounded by including it as a covariate in the subsequent multivariable models presented (Supplementary Fig. 1). Specifically, there was no prior evidence that TMB is significantly associated with ICB response in ccRCC; however, TMB is colinear with driver genes in a small gene panel enriched for cancer genes (e.g. *PBRM1*). Multicollinearity reduces precision of estimated coefficients, which weakens the statistical power of a regression model, and thus one is unable to fully trust the p-values to identify independent variables that are statistically significant. The MSK study then introduce other confounders into the model by combining single agent and dual anti-PD-1/CTLA4 immunotherapies, even though the clinical impact of these drugs in ccRCC may be distinct (19). After removing TMB from their multivariable model, *PBRM1* LOF remains non-statistically significant, although with consistent trends (Cox PH, p = 0.15, HR = 0.75, 95% CI = 0.51 - 1.10). Therefore, statistical issues in TMB analysis and cohort heterogeneity confounded the interpretation of the TTF multivariable model presented in this study. After correction, the multivariable analysis remained underpowered, and while not statistically significant for *PBRM1* LOF, it yielded an association with improved TTF that was consistent with the univariable analysis.

Broadly, we conclude that ICB-associated molecular analyses should explicitly distinguish between two goals: 1) Discovering associations warranting further translational and biological investigation, and 2) Developing predictive biomarkers for immediate clinical use in specific clinical settings. This study provides further evidence in support of our prior findings specifically regarding *PBRM1* LOF associating with single-agent ICB response, which has biological plausibility given preclinical studies of *PBRM1* loss and tumor-immune interactions (1, 22-24) that has prompted further translational investigation. We submit that this study, given the cohort size, p-values close to the threshold used for statistical significance in the setting of modest effect size, technical and statistical anomalies, varying patient populations, and different outcome measurements, does not warrant dismissing a potential biological role of *PBRM1* LOF in ccRCC tumor immunology contexts.

Distinct from direct evaluation of the association of *PBRM1* LOF with benefit from ICI monotherapy, further functional and immunological characterization of other mutations and mutation classes (i.e. missense) in chromatin regulator genes like *PBRM1* using saturation mutagenesis assays may provide more granularity regarding their biological roles. Multiple studies have demonstrated distinct and often subtle functional effects for different missense mutations in different *PBRM1* bromodomains and other mSWI/SNF members, which is further supported by transcriptional differences between LOF, non-LOF (missense or in-frame), and wild-type classes and the absence of specific bromodomain mutation clusters (rather a general increased mutation rate in *PBRM1* exons) (Supplementary Fig. 2) (25-29). As with other tumor suppressors (30, 31), saturation assays could further inform diverse biological effects for *PBRM1* missense mutations in different exonic bromodomains, and potentially their differential impact on tumor-immune interactions, that may not be discriminated by *in silico* or immunohistochemistry techniques (which does not always correlate with genetic loss-of-function) (12, 32).

Most importantly, as we emphasized in prior immunogenomic correlative studies (1, 16, 33, 34), we again strongly caution against overinterpretation of this biological association (among many others) as simultaneously establishing predictive biomarker status in diverse ccRCC clinical contexts or other diseases. For instance, *PBRM1* has also been associated with many other tumor-intrinsic processes (e.g. hypoxia, *TP53* interactions) (25, 35), as well as to VEGF TKI monotherapy response in some clinical settings but not others (4, 5, 36). Since VEGF TKIs and ICB combinations are now a standard of care, and these therapies have numerous complex effects on the already intricate tumor and immune microenvironment interactions, it is unlikely that *PBRM1* LOF (or any single molecular feature, compared to composite transcriptional programs that capture the biological complexity of this context) has strong clinical biomarker relevance in that combination therapy context. Dedicated translational analyses from randomized clinical trials representing specific clinical contexts are necessary to elucidate these factors.

Clinical and molecular data linked to functional studies can invoke new biology but often do not translate to predictive biomarkers. These biological observations may only capture one component of the complex tumor-immune-stromal processes that are perturbed by ICBs and are very unlikely to be driven exclusively by a single molecular event. For the ICB predictive biomarker field, TMB best illustrates this challenge, as it has a biological rationale linked to neoantigens and clinico-genomic associations in specific settings, but has no relevance as a predictive biomarker in ccRCC and may have limited performance as a singleton predictive biomarker in diseases where TMB is established as a genomic correlate of response after considering other molecular and clinical variables (37).

## METHODS

### Survival Analysis

To determine if there are significant differences between the survival curves of two or more groups we used the log-rank test from the survival R package. To determine the hazard ratios and significance of covariates on survival outcomes we performed Cox proportional hazard analysis using the survival R package. To generate the TMB tertile groups for survival analysis, we determined the TMB tertiles from the entire cohort, followed by filtering to only non-LOF and WT *PBRM1* patients.

### Power analysis (simulations)

To perform simulations for power analysis two assumptions were made: (1) The proportion of *PBRM1* mutation statuses (e.g. LOF, non-LOF, and WT) in the authors cohort (n = 189) approximates the relative proportion of these events in ccRCC patient populations. Specifically, the frequency of *PBRM1* LOF mutations in ccRCC patient populations is 32%. (2) The hazard ratios from performing Cox proportional hazard analysis on the originating cohort are the postulated hazard ratios for *PBRM1* LOF mutations in this cancer type and therapeutic settings (e.g. overall, first-line, second-line). To determine the power to detect a significant hazards ratio for *PBRM1* LOF mutations we used the lifelines python package.

### Log transformation of TMB

A total of 7 samples had a TMB of 0. To account for these samples during log transformation, we added a constant 0.5 to the TMB values. For reference, the lowest non-zero TMB observed in the cohort was 0.9 mutations per megabase.

## Data Availability

All data (as provided by the corresponding author of the initiating study) and code written to perform the analyses presented herein is available at: https://github.com/vanallenlab/PBRM1_MSK_RCC_analysis

---

**Supplementary Figure 1:**
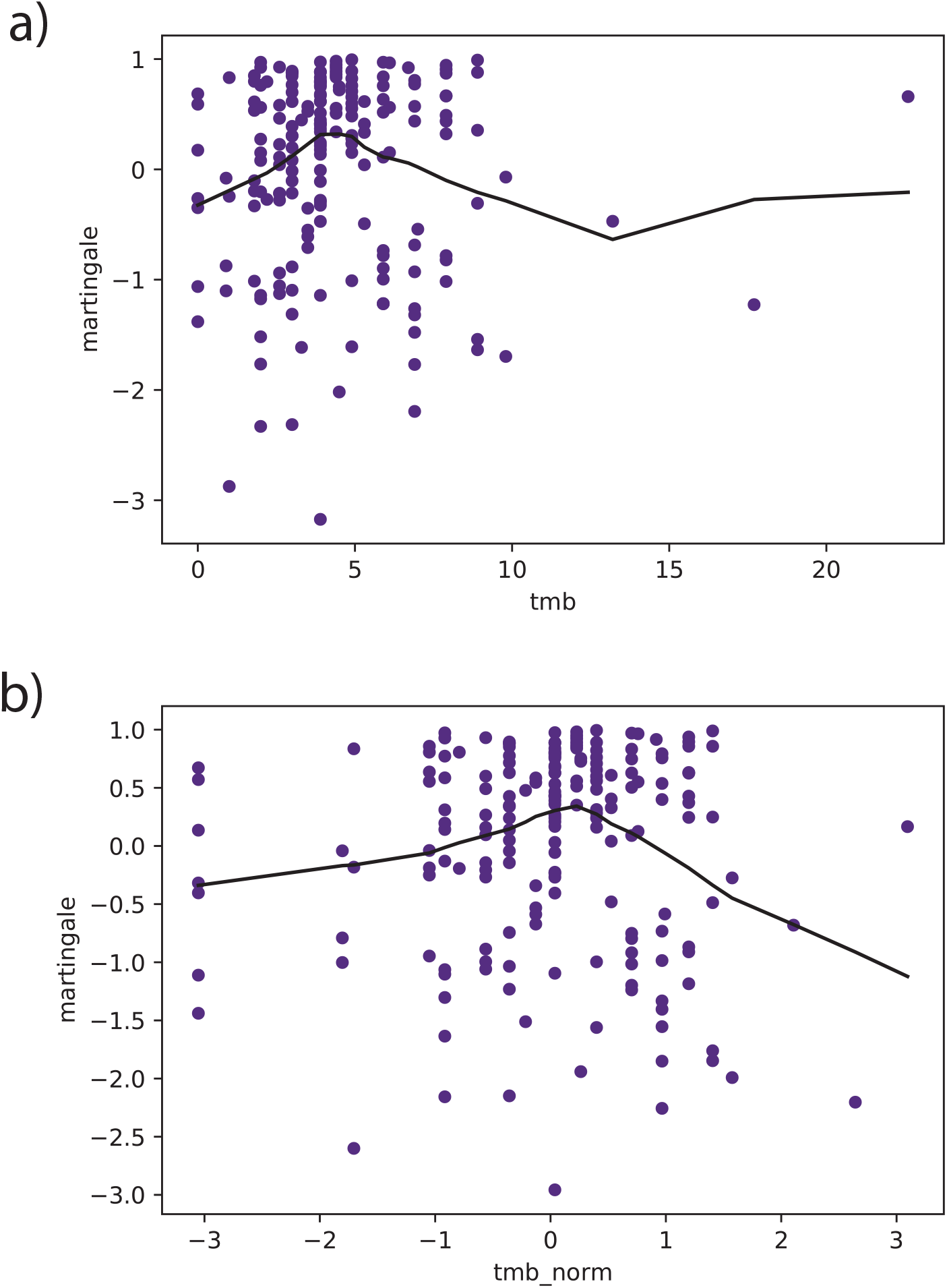
TMB is nonlinear in the originating study data. The martingale residuals for both **a)** TMB and **b)** log-normalized TMB are nonlinear, which violates the Cox proportional hazards model assumption for continuous variables, and suggests that the TMB covariate is not properly fit to the data.

**Supplementary Figure 2:**
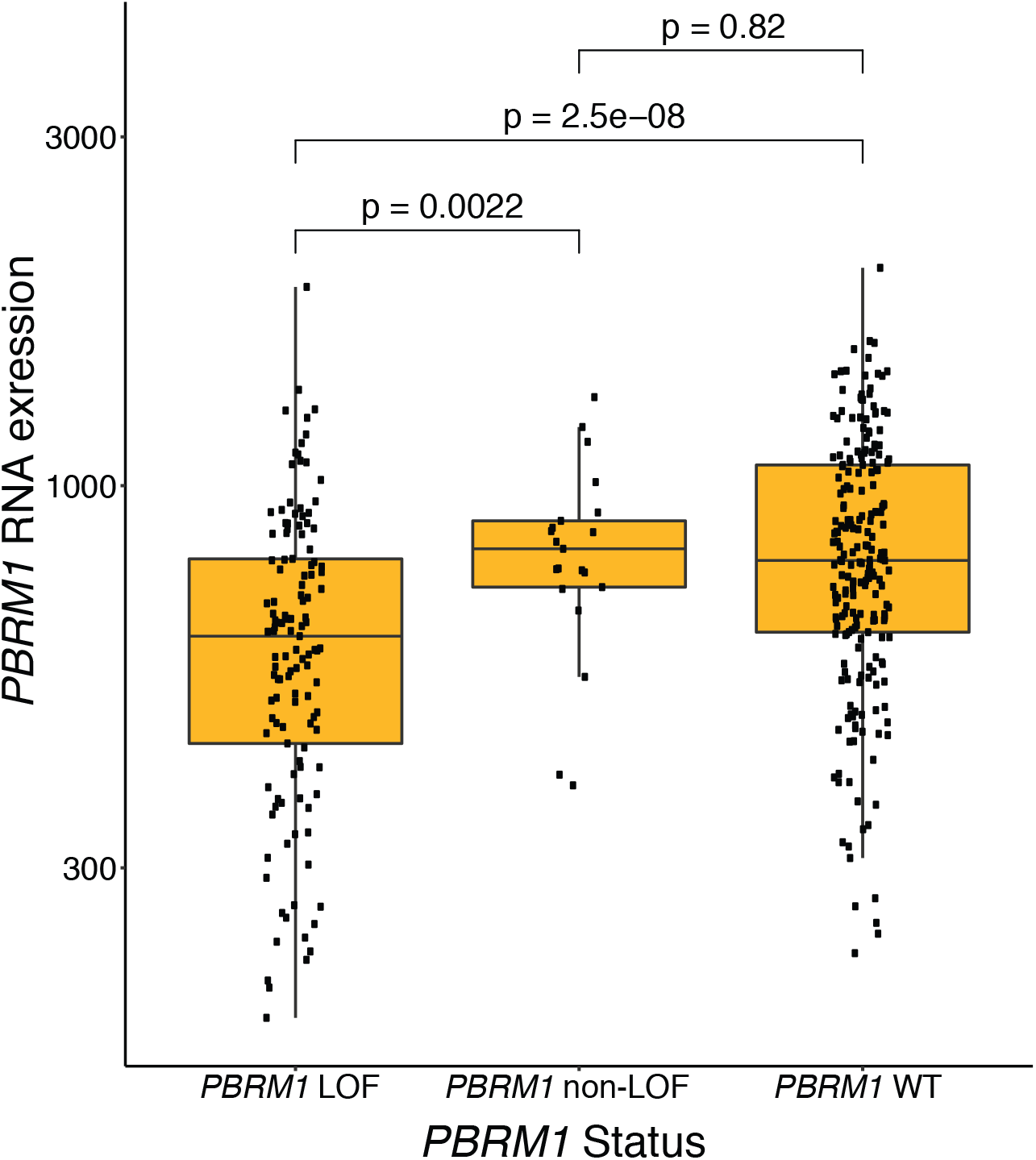
*PBRM1* expression differences by *PBRM1* mutation status. There is no difference in *PBRM1* expression between tumors that are *PBRM1* WT or have non-LOF mutations (e.g. missense and in-frame indels) in *PBRM1*. Conversely, tumors with putative LOF mutations (including splice-site) in *PBRM1* have significantly lower *PBRM1* expression compared to *PBRM1* WT tumors (p = 2.5 x 10^−8^) and tumors with non-LOF mutations (p = 0.0022) in *PBRM1*.

## Notes

### Competing Interest Statement

Advisory/Consulting: Tango Therapeutics, Genome Medical, Invitae, Enara Bio, Janssen, Manifold Bio, Monte Rosa
Research support: Novartis, BMS
Equity: Tango Therapeutics, Genome Medical, Syapse, Enara Bio, Manifold Bio, Microsoft, Monte Rosa
Travel reimbursement: Roche/Genentech
Patents: Institutional patents filed on chromatin mutations and immunotherapy response, and methods for clinical interpretation

### Funding Statement

No external funding was received for this study

### Author Declarations

This study is reanalysis of publicly available data

## REFERENCES

1. Miao D, Margolis CA, Gao W, Voss MH, Li W, Martini DJ, Norton C, Bosse D, Wankowicz SM, Cullen D, Horak C, Wind-Rotolo M, Tracy A, Giannakis M, Hodi FS, Drake CG, Ball MW, Allaf ME, Snyder A, Hellmann MD, Ho T, Motzer RJ, Signoretti S, Kaelin WG, Jr., Choueiri TK, Van Allen EM. Genomic correlates of response to immune checkpoint therapies in clear cell renal cell carcinoma. Science. 2018. Epub 2018/01/06. doi: 10.1126/science.aan5951. PubMed PMID: 29301960.

2. Braun DA, Hou Y, Bakouny Z, Ficial M, Sant’ Angelo M, Forman J, Ross-Macdonald P, Berger AC, Jegede OA, Elagina L, Steinharter J, Sun M, Wind-Rotolo M, Pignon JC, Cherniack AD, Lichtenstein L, Neuberg D, Catalano P, Freeman GJ, Sharpe AH, McDermott DF, Van Allen EM, Signoretti S, Wu CJ, Shukla SA, Choueiri TK. Interplay of somatic alterations and immune infiltration modulates response to PD-1 blockade in advanced clear cell renal cell carcinoma. Nature medicine. 2020;26(6):909–18. Epub 2020/05/31. doi: 10.1038/s41591-020-0839-y. PubMed PMID: 32472114.

3. Braun DA, Ishii Y, Walsh AM, Van Allen EM, Wu CJ, Shukla SA, Choueiri TK. Clinical Validation of PBRM1 Alterations as a Marker of Immune Checkpoint Inhibitor Response in Renal Cell Carcinoma. JAMA Oncol. 2019. Epub 2019/09/06. doi: 10.1001/jamaoncol.2019.3158. PubMed PMID: 31486842; PMCID: PMC6735411.

4. McDermott DF, Huseni MA, Atkins MB, Motzer RJ, Rini BI, Escudier B, Fong L, Joseph RW, Pal SK, Reeves JA, Sznol M, Hainsworth J, Rathmell WK, Stadler WM, Hutson T, Gore ME, Ravaud A, Bracarda S, Suarez C, Danielli R, Gruenwald V, Choueiri TK, Nickles D, Jhunjhunwala S, Piault-Louis E, Thobhani A, Qiu J, Chen DS, Hegde PS, Schiff C, Fine GD, Powles T. Clinical activity and molecular correlates of response to atezolizumab alone or in combination with bevacizumab versus sunitinib in renal cell carcinoma. Nature medicine. 2018;24(6):749–57. Epub 2018/06/06. doi: 10.1038/s41591-018-0053-3. PubMed PMID: 29867230; PMCID: PMC6721896.

5. Motzer RJ, Robbins PB, Powles T, Albiges L, Haanen JB, Larkin J, Mu XJ, Ching KA, Uemura M, Pal SK, Alekseev B, Gravis G, Campbell MT, Penkov K, Lee JL, Hariharan S, Wang X, Zhang W, Wang J, Chudnovsky A, di Pietro A, Donahue AC, Choueiri TK. Avelumab plus axitinib versus sunitinib in advanced renal cell carcinoma: biomarker analysis of the phase 3 JAVELIN Renal 101 trial. Nature medicine. 2020. Epub 2020/09/09. doi: 10.1038/s41591-020-1044-8. PubMed PMID: 32895571.

6. Hakimi AA, Attalla K, DiNatale RG, Ostrovnaya I, Flynn J, Blum KA, Ged Y, Hoen D, Kendall SM, Reznik E, Bowman A, Hwee J, Fong CJ, Kuo F, Voss MH, Chan TA, Motzer RJ. A pan-cancer analysis of PBAF complex mutations and their association with immunotherapy response. Nature communications. 2020;11(1):4168. Epub 2020/08/21. doi: 10.1038/s41467-020-17965-0. PubMed PMID: 32820162; PMCID: PMC7441387.

7. Ged Y, Chaim JL, DiNatale RG, Knezevic A, Kotecha RR, Carlo MI, Lee CH, Foster A, Feldman DR, Teo MY, Iyer G, Chan T, Patil S, Motzer RJ, Hakimi AA, Voss MH. DNA damage repair pathway alterations in metastatic clear cell renal cell carcinoma and implications on systemic therapy. J Immunother Cancer. 2020;8(1). Epub 2020/06/24. doi: 10.1136/jitc-2019-000230. PubMed PMID: 32571992; PMCID: PMC7311069.

8. Ma J, Setton J, Morris L, Albornoz PB, Barker C, Lok BH, Sherman E, Katabi N, Beal K, Ganly I, Powell SN, Lee N, Chan TA, Riaz N. Genomic analysis of exceptional responders to radiotherapy reveals somatic mutations in ATM. Oncotarget. 2017;8(6):10312–23. Epub 2017/01/06. doi: 10.18632/oncotarget.14400. PubMed PMID: 28055970; PMCID: PMC5354661.

9. Yaeger R, Chatila WK, Lipsyc MD, Hechtman JF, Cercek A, Sanchez-Vega F, Jayakumaran G, Middha S, Zehir A, Donoghue MTA, You D, Viale A, Kemeny N, Segal NH, Stadler ZK, Varghese AM, Kundra R, Gao J, Syed A, Hyman DM, Vakiani E, Rosen N, Taylor BS, Ladanyi M, Berger MF, Solit DB, Shia J, Saltz L, Schultz N. Clinical Sequencing Defines the Genomic Landscape of Metastatic Colorectal Cancer. Cancer cell. 2018;33(1):125–36 e3. Epub 2018/01/10. doi: 10.1016/j.ccell.2017.12.004. PubMed PMID: 29316426; PMCID: PMC5765991.

10. Teo MY, Seier K, Ostrovnaya I, Regazzi AM, Kania BE, Moran MM, Cipolla CK, Bluth MJ, Chaim J, Al-Ahmadie H, Snyder A, Carlo MI, Solit DB, Berger MF, Funt S, Wolchok JD, Iyer G, Bajorin DF, Callahan MK, Rosenberg JE. Alterations in DNA Damage Response and Repair Genes as Potential Marker of Clinical Benefit From PD-1/PD-L1 Blockade in Advanced Urothelial Cancers. Journal of clinical oncology : official journal of the American Society of Clinical Oncology. 2018;36(17):1685–94. Epub 2018/03/01. doi: 10.1200/JCO.2017.75.7740. PubMed PMID: 29489427; PMCID: PMC6366295.

11. Jayasinghe RG, Cao S, Gao Q, Wendl MC, Vo NS, Reynolds SM, Zhao Y, Climente-Gonzalez H, Chai S, Wang F, Varghese R, Huang M, Liang WW, Wyczalkowski MA, Sengupta S, Li Z, Payne SH, Fenyo D, Miner JH, Walter MJ, Cancer Genome Atlas Research N, Vincent B, Eyras E, Chen K, Shmulevich I, Chen F, Ding L. Systematic Analysis of Splice-Site-Creating Mutations in Cancer. Cell reports. 2018;23(1):270–81 e3. Epub 2018/04/05. doi: 10.1016/j.celrep.2018.03.052. PubMed PMID: 29617666; PMCID: PMC6055527.

12. Bihr S, Ohashi R, Moore AL, Ruschoff JH, Beisel C, Hermanns T, Mischo A, Corro C, Beyer J, Beerenwinkel N, Moch H, Schraml P. Expression and Mutation Patterns of PBRM1, BAP1 and SETD2 Mirror Specific Evolutionary Subtypes in Clear Cell Renal Cell Carcinoma. Neoplasia. 2019;21(2):247–56. Epub 2019/01/20. doi: 10.1016/j.neo.2018.12.006. PubMed PMID: 30660076; PMCID: PMC6355619.

13. It’s time to talk about ditching statistical significance. Nature. 2019;567(7748):283. Epub 2019/03/22. doi: 10.1038/d41586-019-00874-8. PubMed PMID: 30894740.

14. Tannir NM, McDermott DF, Escudier B, Hammers HJ, Aren OR, Plimack ER, Barthelemy P, Neiman V, George S, Porta C, Powles T, Donskov F, Grimm M-O, Amin A, Tykodi SS, Tomita Y, Rini BI, McHenry MB, Saggi SS, Motzer RJ. Overall survival and independent review of response in CheckMate 214 with 42-month follow-up: First-line nivolumab + ipilimumab (N+I) versus sunitinib (S) in patients (pts) with advanced renal cell carcinoma (aRCC). Journal of Clinical Oncology. 2020;38(6_suppl):609-. doi: 10.1200/JCO.2020.38.6_suppl.609.

15. Motzer RJ, Escudier B, McDermott DF, George S, Hammers HJ, Srinivas S, Tykodi SS, Sosman JA, Procopio G, Plimack ER, Castellano D, Choueiri TK, Gurney H, Donskov F, Bono P, Wagstaff J, Gauler TC, Ueda T, Tomita Y, Schutz FA, Kollmannsberger C, Larkin J, Ravaud A, Simon JS, Xu LA, Waxman IM, Sharma P, CheckMate I. Nivolumab versus Everolimus in Advanced Renal-Cell Carcinoma. The New England journal of medicine. 2015;373(19):1803–13. Epub 2015/09/26. doi: 10.1056/NEJMoa1510665. PubMed PMID: 26406148; PMCID: PMC5719487.

16. Abou Alaiwi S, Nassar AH, Xie W, Bakouny Z, Berchuck JE, Braun DA, Baca SC, Nuzzo PV, Flippot R, Mouhieddine TH, Spurr LF, Li YY, Li T, Flaifel A, Steinharter JA, Margolis CA, Vokes NI, D. H, Shukla SA, Cherniack AD, Sonpavde G, Haddad RI, Awad MM, Giannakis M, Hodi FS, Liu XS, Signoretti S, Kadoch C, Freedman ML, Kwiatkowski DJ, Van Allen EM, Choueiri TK. Mammalian SWI/SNF Complex Genomic Alterations and Immune Checkpoint Blockade in Solid Tumors. Cancer immunology research. 2020;8(8):1075–84. Epub 2020/04/24. doi: 10.1158/2326-6066.CIR-19-0866. PubMed PMID: 32321774; PMCID: PMC7415546.

17. Yao L, Fu Y, Mohiyuddin M, Lam HYK. ecTMB: a robust method to estimate and classify tumor mutational burden. Scientific reports. 2020;10(1):4983. Epub 2020/03/20. doi: 10.1038/s41598-020-61575-1. PubMed PMID: 32188929; PMCID: PMC7080796.

18. Chalmers ZR, Connelly CF, Fabrizio D, Gay L, Ali SM, Ennis R, Schrock A, Campbell B, Shlien A, Chmielecki J, Huang F, He Y, Sun J, Tabori U, Kennedy M, Lieber DS, Roels S, White J, Otto GA, Ross JS, Garraway L, Miller VA, Stephens PJ, Frampton GM. Analysis of 100,000 human cancer genomes reveals the landscape of tumor mutational burden. Genome medicine. 2017;9(1):34. Epub 2017/04/20. doi: 10.1186/s13073-017-0424-2. PubMed PMID: 28420421; PMCID: PMC5395719.

19. McKay RR, Bosse D, Choueiri TK. Evolving Systemic Treatment Landscape for Patients With Advanced Renal Cell Carcinoma. Journal of clinical oncology : official journal of the American Society of Clinical Oncology. 2018:JCO2018790253. Epub 2018/10/30. doi: 10.1200/JCO.2018.79.0253. PubMed PMID: 30372392.

20. Samstein RM, Lee CH, Shoushtari AN, Hellmann MD, Shen R, Janjigian YY, Barron DA, Zehir A, Jordan EJ, Omuro A, Kaley TJ, Kendall SM, Motzer RJ, Hakimi AA, Voss MH, Russo P, Rosenberg J, Iyer G, Bochner BH, Bajorin DF, Al-Ahmadie HA, Chaft JE, Rudin CM, Riely GJ, Baxi S, Ho AL, Wong RJ, Pfister DG, Wolchok JD, Barker CA, Gutin PH, Brennan CW, Tabar V, Mellinghoff IK, DeAngelis LM, Ariyan CE, Lee N, Tap WD, Gounder MM, D’Angelo SP, Saltz L, Stadler ZK, Scher HI, Baselga J, Razavi P, Klebanoff CA, Yaeger R, Segal NH, Ku GY, DeMatteo RP, Ladanyi M, Rizvi NA, Berger MF, Riaz N, Solit DB, Chan TA, Morris LGT. Tumor mutational load predicts survival after immunotherapy across multiple cancer types. Nature genetics. 2019;51(2):202–6. Epub 2019/01/16. doi: 10.1038/s41588-018-0312-8. PubMed PMID: 30643254; PMCID: PMC6365097.

21. Gurjao C, Tsukrov D, Imakaev M, Luquette LJ, Mirny LA. Limited evidence of tumour mutational burden as a biomarker of response to immunotherapy. bioRxiv. 2020:2020.09.03.260265. doi: 10.1101/2020.09.03.260265.

22. Pan D, Kobayashi A, Jiang P, Ferrari de Andrade L, Tay RE, Luoma A, Tsoucas D, Qiu X, Lim K, Rao P, Long HW, Yuan GC, Doench J, Brown M, Liu S, Wucherpfennig KW. A major chromatin regulator determines resistance of tumor cells to T cell-mediated killing. Science. 2018. Epub 2018/01/06. doi: 10.1126/science.aao1710. PubMed PMID: 29301958.

23. Canadas I, Thummalapalli R, Kim JW, Kitajima S, Jenkins RW, Christensen CL, Campisi M, Kuang Y, Zhang Y, Gjini E, Zhang G, Tian T, Sen DR, Miao D, Imamura Y, Thai T, Piel B, Terai H, Aref AR, Hagan T, Koyama S, Watanabe M, Baba H, Adeni AE, Lydon CA, Tamayo P, Wei Z, Herlyn M, Barbie TU, Uppaluri R, Sholl LM, Sicinska E, Sands J, Rodig S, Wong KK, Paweletz CP, Watanabe H, Barbie DA. Tumor innate immunity primed by specific interferon-stimulated endogenous retroviruses. Nature medicine. 2018;24(8):1143–50. Epub 2018/07/25. doi: 10.1038/s41591-018-0116-5. PubMed PMID: 30038220; PMCID: PMC6082722.

24. Liao L, Liu ZZ, Langbein L, Cai W, Cho EA, Na J, Niu X, Jiang W, Zhong Z, Cai WL, Jagannathan G, Dulaimi E, Testa JR, Uzzo RG, Wang Y, Stark GR, Sun J, Peiper S, Xu Y, Yan Q, Yang H. Multiple tumor suppressors regulate a HIF-dependent negative feedback loop via ISGF3 in human clear cell renal cancer. eLife. 2018;7. Epub 2018/10/26. doi: 10.7554/eLife.37925. PubMed PMID: 30355451; PMCID: PMC6234029.

25. Cai W, Su L, Liao L, Liu ZZ, Langbein L, Dulaimi E, Testa JR, Uzzo RG, Zhong Z, Jiang W, Yan Q, Zhang Q, Yang H. PBRM1 acts as a p53 lysine-acetylation reader to suppress renal tumor growth. Nature communications. 2019;10(1):5800. Epub 2019/12/22. doi: 10.1038/s41467-019-13608-1. PubMed PMID: 31863007; PMCID: PMC6925188.

26. Schoenfeld AJ, Bandlamudi C, Lavery JA, Montecalvo J, Namakydoust A, Rizvi H, Egger J, Concepcion CP, Paul S, Arcila ME, Daneshbod Y, Chang J, Sauter JL, Beras A, Ladanyi M, Jacks T, Rudin CM, Taylor BS, Donoghue MTA, Heller G, Hellmann MD, Rekhtman N, Riely GJ. The Genomic Landscape of SMARCA4 Alterations and Associations with Outcomes in Patients with Lung Cancer. Clinical cancer research : an official journal of the American Association for Cancer Research. 2020. Epub 2020/07/28. doi: 10.1158/1078-0432.CCR-20-1825. PubMed PMID: 32709715.

27. Slaughter MJ, Shanle EK, McFadden AW, Hollis ES, Suttle LE, Strahl BD, Davis IJ. PBRM1 bromodomains variably influence nucleosome interactions and cellular function. The Journal of biological chemistry. 2018;293(35):13592–603. Epub 2018/07/11. doi: 10.1074/jbc.RA118.003381. PubMed PMID: 29986887; PMCID: PMC6120218.

28. Porter EG, Dykhuizen EC. Individual Bromodomains of Polybromo-1 Contribute to Chromatin Association and Tumor Suppression in Clear Cell Renal Carcinoma. The Journal of biological chemistry. 2017;292(7):2601–10. Epub 2017/01/06. doi: 10.1074/jbc.M116.746875. PubMed PMID: 28053089; PMCID: PMC5314159.

29. Mashtalir N, Suzuki H, Farrell DP, Sankar A, Luo J, Filipovski M, D’Avino AR, St Pierre R, Valencia AM, Onikubo T, Roeder RG, Han Y, He Y, Ranish JA, DiMaio F, Walz T, Kadoch C. A Structural Model of the Endogenous Human BAF Complex Informs Disease Mechanisms. Cell. 2020. Epub 2020/10/15. doi: 10.1016/j.cell.2020.09.051. PubMed PMID: 33053319.

30. Findlay GM, Daza RM, Martin B, Zhang MD, Leith AP, Gasperini M, Janizek JD, Huang X, Starita LM, Shendure J. Accurate classification of BRCA1 variants with saturation genome editing. Nature. 2018;562(7726):217–22. Epub 2018/09/14. doi: 10.1038/s41586-018-0461-z. PubMed PMID: 30209399; PMCID: PMC6181777.

31. Giacomelli AO, Yang X, Lintner RE, McFarland JM, Duby M, Kim J, Howard TP, Takeda DY, Ly SH, Kim E, Gannon HS, Hurhula B, Sharpe T, Goodale A, Fritchman B, Steelman S, Vazquez F, Tsherniak A, Aguirre AJ, Doench JG, Piccioni F, Roberts CWM, Meyerson M, Getz G, Johannessen CM, Root DE, Hahn WC. Mutational processes shape the landscape of TP53 mutations in human cancer. Nature genetics. 2018;50(10):1381–7. Epub 2018/09/19. doi: 10.1038/s41588-018-0204-y. PubMed PMID: 30224644; PMCID: PMC6168352.

32. Pena-Llopis S, Vega-Rubin-de-Celis S, Liao A, Leng N, Pavia-Jimenez A, Wang S, Yamasaki T, Zhrebker L, Sivanand S, Spence P, Kinch L, Hambuch T, Jain S, Lotan Y, Margulis V, Sagalowsky AI, Summerour PB, Kabbani W, Wong SW, Grishin N, Laurent M, Xie XJ, Haudenschild CD, Ross MT, Bentley DR, Kapur P, Brugarolas J. BAP1 loss defines a new class of renal cell carcinoma. Nature genetics. 2012;44(7):751–9. Epub 2012/06/12. doi: 10.1038/ng.2323. PubMed PMID: 22683710; PMCID: PMC3788680.

33. Miao D, Margolis CA, Vokes NI, Liu D, Taylor-Weiner A, Wankowicz SM, Adeegbe D, Keliher D, Schilling B, Tracy A, Manos M, Chau NG, Hanna GJ, Polak P, Rodig SJ, Signoretti S, Sholl LM, Engelman JA, Getz G, Janne PA, Haddad RI, Choueiri TK, Barbie DA, Haq R, Awad MM, Schadendorf D, Hodi FS, Bellmunt J, Wong KK, Hammerman P, Van Allen EM. Genomic correlates of response to immune checkpoint blockade in microsatellite-stable solid tumors. Nature genetics. 2018;50(9):1271–81. Epub 2018/08/29. doi: 10.1038/s41588-018-0200-2. PubMed PMID: 30150660; PMCID: PMC6119118.

34. Van Allen EM, Miao D, Schilling B, Shukla SA, Blank C, Zimmer L, Sucker A, Hillen U, Foppen MH, Goldinger SM, Utikal J, Hassel JC, Weide B, Kaehler KC, Loquai C, Mohr P, Gutzmer R, Dummer R, Gabriel S, Wu CJ, Schadendorf D, Garraway LA. Genomic correlates of response to CTLA-4 blockade in metastatic melanoma. Science. 2015;350(6257):207–11. doi: 10.1126/science.aad0095. PubMed PMID: 26359337.

35. Gao W, Li W, Xiao T, Liu XS, Kaelin WG, Jr. Inactivation of the PBRM1 tumor suppressor gene amplifies the HIF-response in VHL-/- clear cell renal carcinoma. Proceedings of the National Academy of Sciences of the United States of America. 2017;114(5):1027–32. Epub 2017/01/14. doi: 10.1073/pnas.1619726114. PubMed PMID: 28082722; PMCID: PMC5293026.

36. Hakimi AA, Voss MH, Kuo F, Sanchez A, Liu M, Nixon BG, Vuong L, Ostrovnaya I, Chen YB, Reuter V, Riaz N, Cheng Y, Patel P, Marker M, Reising A, Li MO, Chan TA, Motzer RJ. Transcriptomic Profiling of the Tumor Microenvironment Reveals Distinct Subgroups of Clear Cell Renal Cell Cancer: Data from a Randomized Phase III Trial. Cancer discovery. 2019;9(4):510–25. Epub 2019/01/10. doi: 10.1158/2159-8290.CD-18-0957. PubMed PMID: 30622105; PMCID: PMC6697163.

37. Liu D, Schilling B, Liu D, Sucker A, Livingstone E, Jerby-Amon L, Zimmer L, Gutzmer R, Satzger I, Loquai C, Grabbe S, Vokes N, Margolis CA, Conway J, He MX, Elmarakeby H, Dietlein F, Miao D, Tracy A, Gogas H, Goldinger SM, Utikal J, Blank CU, Rauschenberg R, von Bubnoff D, Krackhardt A, Weide B, Haferkamp S, Kiecker F, Izar B, Garraway L, Regev A, Flaherty K, Paschen A, Van Allen EM, Schadendorf D. Integrative molecular and clinical modeling of clinical outcomes to PD1 blockade in patients with metastatic melanoma. Nature medicine. 2019;25(12):1916–27. Epub 2019/12/04. doi: 10.1038/s41591-019-0654-5. PubMed PMID: 31792460; PMCID: PMC6898788.

